# The Need for Developing Consensus-Based Guidelines in Assessing Parkinson’s Disease Laterality

**DOI:** 10.1101/2025.07.24.25332091

**Authors:** Emily Houston, Sean Boyd, Vivikta Iyer, James Boyd, Deepak Gupta

**Affiliations:** Binter Center for Parkinson’s Disease & Movement Disorders, University of Vermont Medical Center; Department of Neurological Sciences, Larner College of Medicine, University of Vermont

**Keywords:** Parkinson’s disease, Laterality, Parkinsonism, Hoehn and Yahr scale (HY), Movement Disorders Society Parkinson’s Disease Rating Scale (MDS-UPDRS)

## Abstract

This study investigated the correlation between Parkinson’s disease laterality, as defined by the Hoehn and Yahr (HY) scale, and appendicular parkinsonism scores from the MDS-UPDRS Part III using the PPMI dataset. While most cases (89.6%) showed congruence, a 10.4% discordance suggests that factors beyond appendicular parkinsonism influence clinical HY staging, highlighting a need for standardized guidelines.

## Background

The Hoehn and Yahr scale (HY) is currently the most widely used staging system for Parkinson’s disease (PD) and is formally a part of the Movement Disorders Society Parkinson’s Disease Rating Scale (MDS-UPDRS).^1, 2^ Transition from HY stage 1 to 2 is considered a key milestone of PD progression and often used as eligibility criterion for early PD clinical trials. The original HY publication defined stage 1 as *“Functional impairment is minimal, but unilateral features of tremor, rigidity, or bradykinesia are evident”* and stage 2 as *“Balance is not yet impaired, but the features in stage 1 become bilateral”*.^1^ The clear defining difference in transitioning from HY 1 to 2 is the progression from unilateral to bilateral signs, in the absence of postural instability.

We aimed to investigate if PD laterality in HY staging would correlate with scores anchored to appendicular parkinsonism (defined as motor features of bradykinesia, rigidity, and/or rest tremor in limbs) in the MDS-UPDRS part III.

## Patients and Methods

We downloaded the Parkinson Progression Marker Initiative (PPMI) dataset on November 12^th^, 2024 ((www.ppmi-info.org/access-data-specimens/download-data), RRID:SCR_006431. We first defined appendicular parkinsonism as unilateral or bilateral based on relevant MDS-UPDRS motor variables having a value of ≥1 on either side or both sides. The motor variables included features of bradykinesia (items 3.4 to 3.8), rigidity (item 3.3), and rest tremor (item 3.17). We then compared PD laterality based on appendicular parkinsonism with HY stage 1 (unilateral) and stage ≥2 (bilateral).

## Results and Discussions

Of the 15,567 PPMI participants, H&Y-based unliteral and bilateral cases were 23.3% and 76.7%, respectively, while appendicular parkinsonism-based unilateral and bilateral cases were 18.3% and 81.7%, respectively.

Upon cross-tabulation of H&Y-based laterality and parkinsonism laterality (table 1), there was a match in 89.6% cases. Of the 10.4% unmatched cases, 7.7% cases were HY-based unilateral despite bilateral appendicular parkinsonism, while 2.7% cases were HY-based bilateral despite unilateral appendicular parkinsonism on MDS-UPDRS part III.

**Table 1:** Cross-tabulation of PD Laterality Based on the HY Stage and Appendicular Parkinsonism.

| PD Laterality<br>(Total N= 15,567) | HY Stage 1 Unilateral<br>N (% of total) | HY Stage 2 Bilateral<br>N (% of total) |
| --- | --- | --- |
| Parkinsonism Unilateral<br>N (% of total) | 2,422 (15.6%) | 421 (2.7%) |
| Parkinsonism Bilateral<br>N (% of total) | 1,207(7.7%) | 11,526(74.0%) |

In the PPMI dataset, most cases demonstrated congruence between HY-based laterality and the appendicular parkinsonism laterality. Our findings indicate that factors other than appendicular parkinsonism might be used by movement disorders specialists in assessing laterality and assigning a HY stage, leading to the 10.4% discordance. For instance, the presence of postural instability despite unilateral appendicular parkinsonism might be graded as HY 3.^3^ Alternatively, a person with PD showing minimal signs of a parkinsonism feature on their “non-affected” side may be categorized as bilateral on the MDS-UPDRS, but their neurologist deems them to be H&Y 1, based on knowledge of their global presentation. Another possibility might be that movement disorders neurologists assess PD as bilateral in H&Y staging due to presence of action tremor on the side unaffected by appendicular parkinsonism. These findings further emphasize the need for developing consensus-based guidelines for assessing laterality of PD in HY stage and in using the MDS-UPDRS in general.

## Data Availability

All data produced in the present study is available on the PPMI website.

https://www.ppmi-info.org/access-data-specimens/download-data

## Acknowledgements

We wish to acknowledge the efforts of the PPMI Investigators, study teams, and especially the participants in generating this large dataset. We are grateful for the opportunity to access and analyze the data, in an effort to contribute meaningful findings to the broad PD community.

D.G., S.B., J.B.,V.I. and E.H. contributed to the conception of the work, and D.G. directly oversaw and mentored S.B. on the analysis. All authors were involved in the interpretation of the results. E.H. wrote the initial draft of the letter, with D.G. and J.B. revising it for intellectual content. The final version was reviewed and approved by all authors.

